# A comprehensive analysis of dominant and recessive parkinsonism genes in REM sleep behavior disorder

**DOI:** 10.1101/2020.03.17.20032664

**Authors:** Kheireddin Mufti, Uladzislau Rudakou, Eric Yu, Jennifer A. Ruskey, Farnaz Asayesh, Sandra B. Laurent, Dan Spiegelman, Isabelle Arnulf, Michele T.M. Hu, Jacques Y. Montplaisir, Jean-François Gagnon, Alex Desautels, Yves Dauvilliers, Gian Luigi Gigli, Mariarosaria Valente, Francesco Janes, Birgit Högl, Ambra Stefani, Evi Holzknecht, Karel Sonka, David Kemlink, Wolfgang Oertel, Annette Janzen, Giuseppe Plazzi, Elena Antelmi, Michela Figorilli, Monica Puligheddu, Brit Mollenhauer, Claudia Trenkwalder, Friederike Sixel-Döring, Valérie Cochen De Cock, Christelle Charley Monaca, Anna Heidbreder, Luigi Ferini-Strambi, Femke Dijkstra, Mineke Viaene, Beatriz Abril, Bradley F. Boeve, Ronald B. Postuma, Guy A. Rouleau, Ziv Gan-Or

## Abstract

**Objective:** To examine the role of autosomal dominant (AD) and recessive (AR) Parkinsonism genes in the risk of isolated rapid-eye-movement (REM) sleep behavior disorder (iRBD).

**Methods:** Ten genes implicated in AD and AR Parkinsonism were fully sequenced using targeted next-generation sequencing in 1,039 iRBD patients and 1,852 controls of European ancestry. These include the AR genes *PRKN, DJ-1 (PARK7), PINK1, VPS13C, ATP13A2, FBXO7* and *PLA2G6*, and the AD genes *LRRK2, GCH1* and *VPS35*. To examine the role of rare heterozygous variants in these genes, burden test and SKAT-O analyses were performed. The contribution of homozygous and compound heterozygous variants was further examined in the AR genes. Copy number variants (CNVs) in *PRKN* were tested in a subset of samples (n=374) using multiplex ligation-dependent probe amplification followed by analysis of all samples using ExomeDepth.

**Results:** We found no association between rare heterozygous variants in the tested genes and risk for iRBD. Several homozygous and compound heterozygous carriers were identified with variants of unknown significance, yet there was no overrepresentation in iRBD patients versus controls.

**Conclusion:** Our results do not support a major role for variants in *PRKN, PARK7, PINK1, VPS13C, ATP13A2, FBXO7, PLA2G6, LRRK2, GCH1* and *VPS35* in the risk of iRBD.

## Introduction

Isolated rapid eye movement (REM)-sleep behavior disorder (iRBD) is a prodromal neurodegenerative disease. More than 80% of iRBD patients diagnosed with video-polysomnography (vPSG) will eventually convert to an overt α-synucleinopathy.^1^ These include mostly Parkinson’s disease (PD) and dementia with Lewy bodies (DLB), and a small minority will convert to multiple system atrophy (MSA).^2^

While not much is known about the genetic background of DLB and MSA, accumulating data from the last two decades have unraveled the role of common and rare genetic variants in PD. Currently, 90 independent risk factors of PD in 78 genetic loci are known, discovered through genome-wide association studies (GWAS).^3^ Other, less common genetic variants, have been implicated in familial forms of PD, including autosomal dominant (AD) inherited variants in genes such as *SNCA, LRRK2, GCH1* and *VPS35*,^4-6^ and autosomal recessive (AR) inherited variants in *PRKN, PINK1* and *PARK7*.^7^ Bi-allelic mutations in other genes, including *ATP13A2, VPS13C, FBXO7* and *PLA2G6* may cause AR atypical syndromes with Parkinsonism,^4,8^ in some of which α-synucleinopathy has also been reported.^9-11^

The genetic background of iRBD has only been studied in recent years, with studies showing that there is no full genetic overlap between the genetic background of iRBD and that of PD or DLB. For example, *GBA* mutations are associated with risk of iRBD, PD and DLB,^2,12^ but pathogenic *LRRK2* mutations seem to be involved only in PD and not in iRBD and DLB.^8,13,14^ *MAPT* and *APOE* variants are important risk factors of PD and DLB, respectively,^15,16^ but both genes are not associated with iRBD.^15,17^ In the *SNCA* locus, there are independent risk variants of PD, DLB and iRBD; specific 3’ variants are associated with PD, and other, independent variants at the 5’ of *SNCA* are associated with iRBD and DLB.^18^ Within the *TMEM175* locus, there are two independent risk factors of PD, but only one of them, the coding polymorphism p.M393T, has also been associated with iRBD.^19^

Thus far, the role of most of the familial PD genes or genes involved in rare forms of atypical parkinsonism has not been studied in iRBD. Here, since *GBA* and *SNCA* have been studied previously,^2,18^ we aimed to thoroughly examine the roles of *PRKN, PINK1, PARK7* (*DJ-1*), *VPS13C, ATP13A2, FBXO7, PLA2G6, LRRK2, GCH1* and *VPS35* in iRBD.

## Methods

### Population

A total of 1,039 unrelated iRBD patients and 1,852 unrelated controls were included in this study, all of European ancestry (confirmed by principal component analysis of GWAS data). Approximately 81% of the patients were male, the mean reported age at onset (AAO) was 60.1 ± 10.5 years and the average age at diagnosis was 65.3 ± 8.7 years. Data on sex and age were available for 1,032 and 1,004 patients, respectively. Among the controls, about 51% were male, and the mean age at sampling was 52.3 ± 14.3 years, age was not available for nine controls. RBD diagnosis was done with video polysomnography according to the ICSD-2/3 criteria (International Classification of Sleep Disorders, version 2 or 3).^20^

### Standard protocol approvals, registrations, and patient consents

All patients signed an informed consent form before entering the study, and the study protocol was approved by the institutional review boards.

### Genetic analysis

The coding sequences and 5’ and 3’ untranslated regions (UTRs) of *PRKN, PINK1, DJ-1, VPS13C, ATP13A2, FBXO7, PLA2G6, LRRK2, GCH1* and *VPS35* were captured using molecular inversion probes (MIPs) designed as previously described,^21^ and the full protocol is available upon request. Details of the MIPs used in the current study are listed in Supplementary Table 1. The library was sequenced on illumina HiSeq 2500\4000 platform at the McGill University and Génome Québec Innovation Centre. Sequencing reads were mapped to the human reference genome (hg19) using the Burrows-Wheeler Aligner.^22^ Post-alignment quality control and variant calling were done using the Genome Analysis Toolkit (GATK, v3.8),^23^ and annotation with ANNOVAR.^24^ The Frequency of each variant was extracted from the Genome Aggregation Database (GnomAD).^25^ We used ClinVar and specific searches on PubMed to examine whether variants that were found in these genes are known or suspected to be pathogenic in PD or atypical parkinsonism.

### Quality control

To perform quality control (QC), we used the PLINK software. We excluded variants with: genotyping rate lower than 90%, deviation from Hardy-Weinberg equilibrium set at *p*=0.001 threshold and when the variant was identified in <25% of the reads for a specific variant. To be included in the analysis, the minimum quality score (QS) was set to 30. Threshold for rate of missingness difference between cases and controls was set at *p*=0.05, and variants below this threshold were removed. Genotyping rate cut-off for individuals was 90%, and individuals with a lower genotyping rate were excluded. After the QC steps, 1,039 patients and 1,852 controls were included in the analysis. Since we aimed to examine the role of variants that cause monogenic PD, only rare variants (minor allele frequency [MAF]<0.01) were included in the analysis. To ensure that we capture high quality variants, we performed analyses for variants with coverage depth of >30X and variants with >50X.

### Data and statistical analysis

We used different approaches to examine the effect of multiple variants on iRBD risk. To examine whether there is a burden of rare (MAF<0.01) heterozygous variants in each of our targeted genes, we used optimized sequence Kernel association test (SKAT-O, R package)^26^ and burden tests for different types of variants: all rare variants, potentially functional rare variants (nonsynonymous, frame-shift, stop-gain and splicing), rare loss-of-function variants (frame-shift, stop-gain and splicing), and rare nonsynonymous variants only. We then examined the association between variants predicted to be pathogenic based on Combined Annotation Dependent Depletion (CADD) score of ≥12.37 (representing the top 2% of potentially deleterious variants) and iRBD. For this analysis, we used burden test (R package SKAT) since the direction of the association was presumed as pathogenic prior to the test. In addition, since copy number variants (CNVs) are frequent in the *PRKN* gene,^27^ we included CNVs when we analyzed the association of *PRKN* variants with iRBD. To call CNVs, we first performed multiplex ligation probe amplification (MLPA, the gold standard for CNV detection in *PRKN*) analysis of 374 samples using the SALSA MLPA Probemix P051 Parkinson mix 1 according to the manufacturer’s instructions (MRC Holland). Then, using the ExomDepth tool,^28^ we determined the ideal parameters for CNV calls using the MIPs data, with sensitivity of 100% and specificity of 97% when compared to the MLPA results. These parameters were subsequently applied to call CNVs from the MIPs data across all iRBD patients and controls. The contribution of homozygous and compound heterozygous variants in all the genes was also examined by comparing the frequencies of the very rare (MAF<0.001) nonsynonymous, splice-site, frame-shift and stop-gain variants between patients and controls. Bonferroni correction for multiple comparisons was applied in all analyses.

### Availability of data and materials

Data used for the analysis is available in the supplementary tables. Anonymized raw data can be shared upon request from any qualified investigator.

## Results

### Quality of coverage

The average coverage of the 10 genes analyzed in this study was >144X for all genes, and the coverage of 8 of the genes was >900X. The per-gene coverage for all 10 genes, although not perfect, is better than the coverage of these specific genes in gnomAD. Supplementary Table 2 details the average coverage and the percentage of nucleotides covered at 20X and 50X for each gene. There were no differences in the coverage across the samples (patients and controls).

### Rare homozygous and compound heterozygous variants are not enriched in iRBD patients

To examine whether homozygous or compound heterozygous variants in our genes of interest may cause iRBD, we compared the carrier frequencies of very rare (MAF <0.001) bi-allelic variants between iRBD patients and controls. Three carriers (one patient and two controls) were identified with homozygous variants across all genes. All three carried homozygous non-coding variants that are not likely to cause a disease: one male patient with AAO of 76 years who carried the *PINK1* variant rs181532922, c.*717T>C at the 3’ UTR of the gene, one female control recruited at age 72 who carried the *DJ-1* rs7534132, an intronic variant, and one control recruited at the age of 26 who carried the *LRRK2* rs72546315 synonymous (p.H275H) variant.

For the analysis of compound heterozygous carriers, since phasing could not be performed, we considered carriers of two rare variants as compound heterozygous carriers, with two exceptions: 1) when variants were physically close and we could determine their phase based on the sequence reads and 2) if the same combination of very rare variants appeared more than once, we assumed that the variants are likely on the same allele. We found a total of 9 patients and controls, presumably compound heterozygous carriers in the studied genes (Table 1). Three affected and three unaffected carriers of compound heterozygous variants in *VPS13C* were identified, with no overrepresentation in iRBD patients (Fisher test, *p*=1).

**Table 1.**
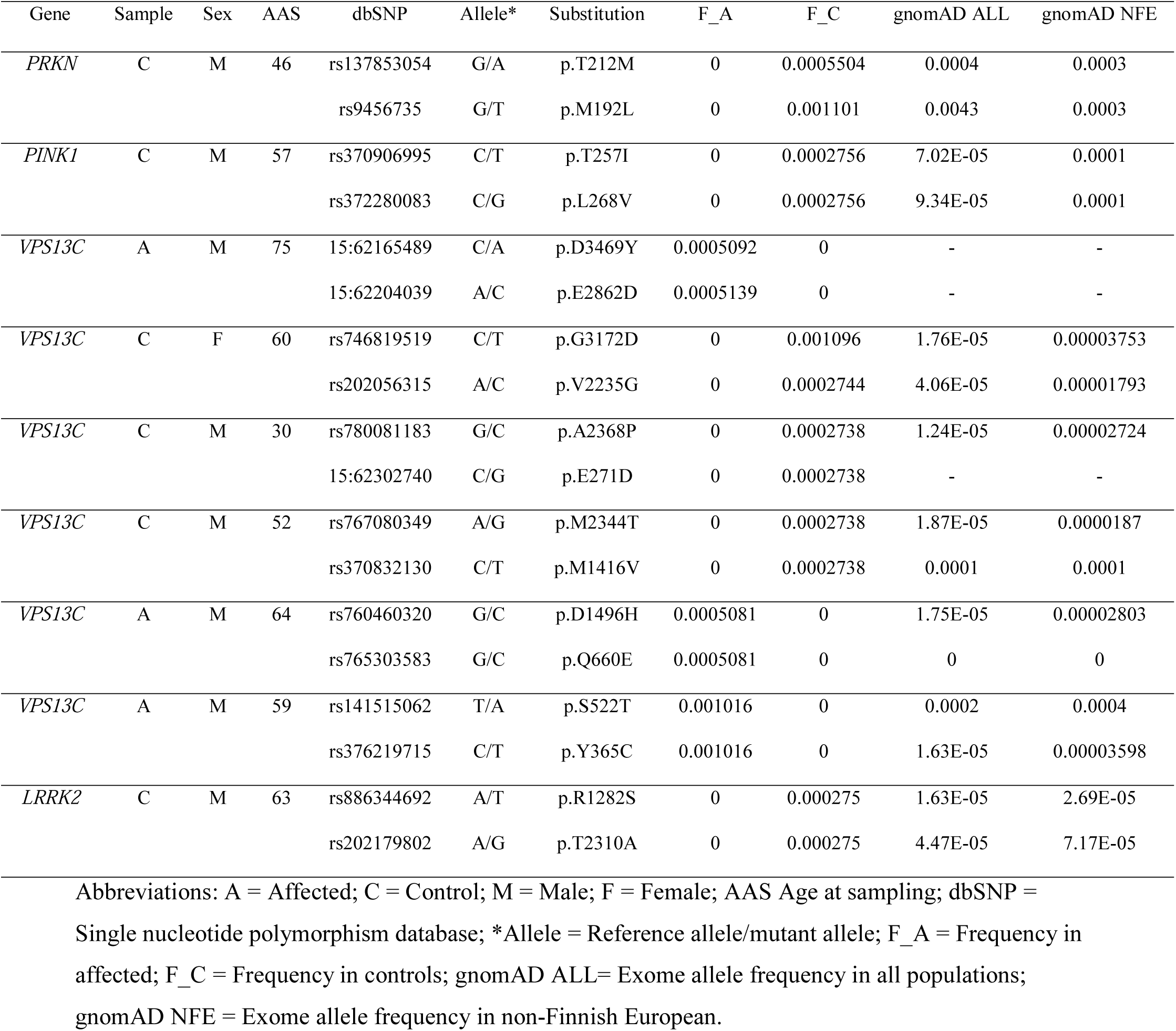
Summary of all samples carrying two nonsynonymous variants detected in the present study

### Rare heterozygous variants are not enriched in any of the studied genes

In order to further study the role of rare (MAF<0.01) heterozygous variants, we performed SKAT-O and burden tests, repeated twice for variants detected at coverage depth of >30X and variants detected at >50X (see methods). All rare heterozygous variants identified in each gene are detailed in supplementary table 3. We performed SKAT-O and burden tests at 5 different levels: all rare variants, all potentially functional variants (nonsynonymous, splice site, frame-shift and stop-gain), loss-of-function variants (frame-shift, stop-gain and splicing), nonsynonymous variants only, and variants with CADD score ≥12.37 (Table 2). The Bonferroni corrected *p* value for statistical significance was set on *p*<0.001. We found no statistically significant association between iRBD and any of the variant types in any of the genes, suggesting that these genes either have no role in iRBD or have a minor role that we could not detect with this sample size. The nominal association between *PARK7* and iRBD in the SKAT-O analysis of rare functional variants is driven by the nonsynonymous variant rs71653622 (p.A179T) which was ∼10 times more frequent in iRBD patients (0.003074) compared to controls (0.000277), but not statistically significant (*p*=0.09, see supplementary table 3). We did not identify any iRBD patient with known biallelic pathogenic variants in *PARK7, PINK1, VPS13C* and *ATP13A2*, or heterozygous pathogenic variants in *LRRK2, GCH1* and *VPS35*. Two controls were found with the pathogenic *LRRK2* p.G2019S variant. We identified 9 (0.86%) iRBD patients and 13 (0.70%, *p*=0.65) controls who were heterozygous carriers of the potentially pathogenic variant p.R275W in *PRKN*, and two additional controls with the *PRKN* p.T240M pathogenic variant. One patient and one control with the pathogenic variant p.R299C in *FBXO7* were also found.

**Table 2.**
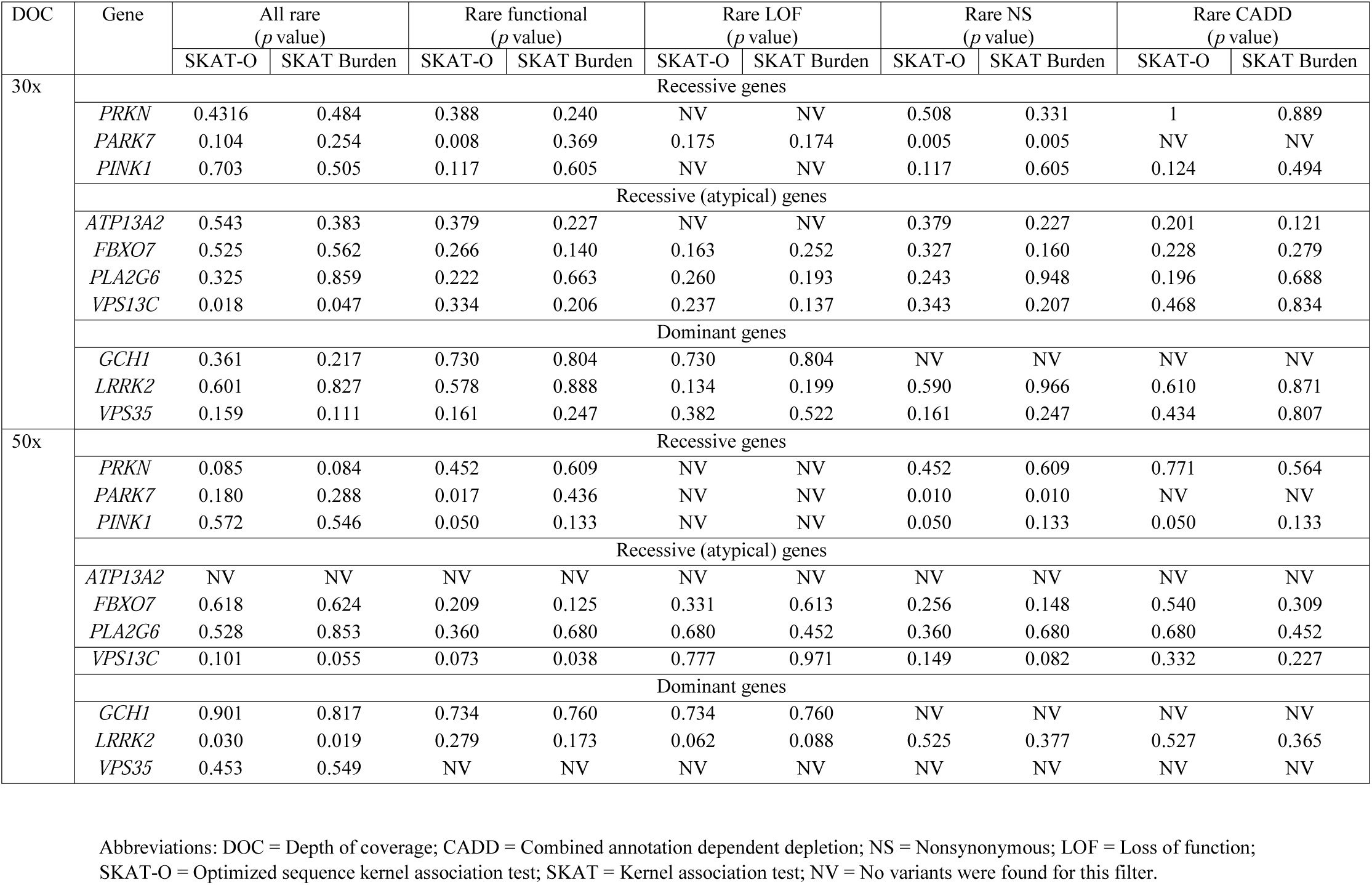
Summary of results from burden analyses of rare heterozygous variants

### Analysis of copy number variants in *PRKN*

We further examined the association between deletions and duplications in *PRKN* and risk for iRBD. Using ExomeDepth, 7 patients (0.7 %) and 17 controls (0.9%, *p*=0.53) were found to carry CNVs in *PRKN*, and none of the patients found to have an additional nonsynonymous variant. Therefore, there were no homozygous or compound heterozygous carriers of rare *PRKN* variants among the iRBD patients. Supplementary table 4 lists all the CNVs found in our cohort.

## Discussion

The present study provides the first large-scale, full sequencing analysis to examine the possible role of the dominant and recessive parkinsonism genes *PRKN, PARK7, PINK1, VPS13C, ATP13A2, FBXO7, PLA2G6, LRRK2, GCH1* and *VPS35* in iRBD. We did not find evidence for association of any of these genes with iRBD. In the recessive genes, there was no over-representation of carriers of homozygous or compound heterozygous variants in iRBD patients, and no single patient with bi-allelic pathogenic variants. In the dominant genes, we did not find any known pathogenic variants in these genes, and SKAT-O and burden analyses did not identify burden of rare heterozygous variants in any of these 10 genes.

Whether heterozygous carriage of mutations in recessive PD or atypical parkinsonism related genes is a risk factor for PD is still controversial.^29^ *PRKN*-associated PD is characterized by pure nigral degeneration without α-synuclein accumulation,^30^ and reports on synucleinopathy and Lewy bodies in *PINK1*-associated PD are inconclusive, as some studies identified Lewy bodies while others did not.^31,32^ Since iRBD is a prodromal synucleinopathy, it is not surprising that we did not identify bi-allelic mutations or burden of heterozygous variants in any of these genes. Of note, 380 (36.5%) of the iRBD cohort had a self-reported AAO <50 years. In the case of iRBD, reported AAO may be especially unreliable, as patients may have had RBD symptoms long before they were noticed by themselves or their bed partners. Therefore, the true percentage of iRBD patients with AAO <50 is likely higher, yet none of the known genes involved in early onset PD seems to be involved in early onset iRBD.

Recently, we have shown that the *SNCA* locus is important in RBD, yet with different and distinct variants that are associated with risk of PD.^18^ In the same study, *SNCA* was fully sequenced and no known PD-causing variants were found in iRBD patients. We and others have previously reported that pathogenic *LRRK2* variants were not identified in smaller cohorts of iRBD,^14^ which was further confirmed in the current study. In addition, several studies of PD patients with and without RBD have shown reduced prevalence of RBD ^33-36^ or reduced scores in RBD questionnaires among *LRRK2* mutation carriers. *VPS35* mutations have not been identified in iRBD in the current study, although pathogenic *VPS35* mutations are generally rare.^37,38^ Altogether, these results provide no evidence that known, well-validated familial gene mutations involved in PD (including *SNCA, LRRK2, VPS35, PRKN, PINK1* and *PARK7*) are also involved in iRBD. *GBA* is the only gene in which strong risk variants associated with PD are also associated with iRBD.^2^

Our study has some limitations. While being the largest genetic study of iRBD to date, it may still be underpowered to detect rare variants in familial PD-related genes. Therefore, our study does not completely rule out the possibility that variants in these genes may lead to iRBD in very rare cases. Another potential limitation of the study design is the earlier age and the different sex distribution in the control population, and the fact that they have not been tested for iRBD. However, since iRBD is not common, found in about 1% of the population,^1^ age would have a minimal or no effect on the results. The differences in sex ratios are less likely to have an effect, since in AD and AR Mendelian diseases, the risk is typically similar for men and women.

To conclude, the lack of association between different PD and Parkinsonism genes may suggest that either iRBD is an entity more affected by environmental factors, or that there are other, yet undetected genes that may be involved in iRBD. To examine these possibilities, larger studies that include carefully collected epidemiological data and more extensive genetic data such as whole-exome or whole-genome sequencing will be required. Our study also suggests that screening for variants in the tested genes will have a very low yield.

## Glossary

*ATP13A2*: Probable cation-transporting ATPase 13A2
AD: Autosomal dominant
AR: Autosomal recessive
CNVs: Copy number variants
*FBXO7*: F-box only protein 7
*GCH1*: GTP cyclohydrolase I
iRBD: Isolated rapid eye movement (REM) sleep behavior disorder
*LRRK2*: Leucine-rich repeat kinase 2
MIPs: Molecular Inversion Probes
NGS: Next generation sequencing
*PARK7*: Parkinson disease protein 7
PD: Parkinson’s disease
*PINK1*: PTEN-induced kinase 1
*PLA2G6*: 85 kDa calcium-independent phospholipase A2
*PRKN*: Parkinson disease protein 2
*VPS13C*: Vacuolar protein sorting-associated protein 13C
*VPS35*: Vacuolar protein sorting-associated protein 35.

## Acknowledgment

This work was supported by the Michael J. Fox Foundation, the Canadian Consortium on Neurodegeneration in Aging (CCNA), Parkinson Canada and the Canada First Research Excellence Fund (CFREF), with funds awarded to McGill University for the Healthy Brains for Healthy Lives (HBHL) program. The Montreal cohort was funded by the Canadian Institutes of Health Research (CIHR) and the W. Garfield Weston Foundation. The Oxford Discovery study is funded by the Monument Trust Discovery Award from Parkinson’s UK and supported by the National Institute for Health Research (NIHR) Oxford Biomedical Research Centre based at Oxford University Hospitals NHS Trust and University of Oxford, the NIHR Clinical Research Network and the Dementias and Neurodegenerative Diseases Research Network (DeNDRoN). JFG holds a Canada Research Chair in Cognitive Decline in Pathological Aging. WO is Hertie Senior Research Professor, supported by the Hertie Foundation. EAF holds a Canada Research Chair (Tier 1) in Parkinson disease. GAR holds a Canada Research Chair in Genetics of the Nervous System and the Wilder Pen field Chair in Neurosciences. ZGO is supported by the Fonds de recherche du Québec–Santé Chercheur-Boursier award and is a Parkinson Canada New Investigator awardee. We thank D. Rochefort, H. Catoire, and V. Zaharieva for their assistance.

## Appendix 1 Authors

| **Name** | **Location** | **Role** | **Contribution** |
| --- | --- | --- | --- |
| **Kheireddin Mufti,**<br>**BPhar** | McGill University, QC, Canada | Author | Designed and conceptualized study;<br>analyzed the data; drafted the manuscript<br>for intellectual content |
| **Uladislau Rudakou,**<br>**BSc** | McGill University, QC, Canada | Author | Analyzed the data; revised the manuscript<br>for intellectual content |
| **Eric Yu, BSc** | McGill University, QC, Canada | Author | Analyzed the data; revised the manuscript<br>for intellectual content |
| **Jennifer A. Ruskey,**<br>**MSc** | McGill University, QC, Canada | Author | Major role in the acquisition of data;<br>revised the manuscript for intellectual<br>content |
| **Farnaz Asayesh, MSc** | McGill University, QC, Canada | Author | Major role in the acquisition of data;<br>revised the manuscript for intellectual<br>content |
| **Sandra B. Laurent,**<br>**BTS** | McGill University, QC, Canada | Author | Major role in the acquisition of data;<br>revised the manuscript for intellectual<br>content |
| **Dan Spiegelman, MSc** | McGill University, QC, Canada | Author | Major role in the acquisition of data;<br>revised the manuscript for intellectual<br>content |
| **Isabelle Arnulf, MD,**<br>**PhD** | Moelle Epinière and Sorbonne<br>Universities, Paris, France | Author | Major role in the acquisition of data;<br>revised the manuscript for intellectual<br>content |
| **Michele T.M. Hu,**<br>**MBBS, FRCP, PhD** | Oxford University, Oxford, UK | Author | Major role in the acquisition of data;<br>revised the manuscript for intellectual<br>content |
| **Jacques Y. Montplaisir, MD, PhD** | Université de Montréal, QC, Canada | Author | Major role in the acquisition of data; revised the manuscript for intellectual content |
| **Jean-François Gagnon, PhD** | Hôpital du Sacré-Cœur de Montréal, QC, Canada | Author | Major role in the acquisition of data; revised the manuscript for intellectual content |
| **Alex Desautels, MD, PhD** | Université de Montréal, QC, Canada | Author | Major role in the acquisition of data; revised the manuscript for intellectual content |
| **Yves Dauvilliers, MD, PhD** | University of Montpellier, Montpellier, France | Author | Major role in the acquisition of data; revised the manuscript for intellectual content |
| **Gian Luigi Gigli, MD** | University Hospital of Udine, Udine, Italy | Author | Major role in the acquisition of data; revised the manuscript for intellectual content |
| **Mariarosaria Valente, MD** | University Hospital of Udine, Udine, Italy | Author | Major role in the acquisition of data; revised the manuscript for intellectual content |
| **Francesco Janes, MD, PhD** | University Hospital of Udine, Udine, Italy | Author | Major role in the acquisition of data; revised the manuscript for intellectual content |
| **Birgit Högl, MD** | Medical University of Innsbruck, Innsbruck, Austria | Author | Major role in the acquisition of data; revised the manuscript for intellectual content |
| **Ambra Stefani, MD** | Medical University of Innsbruck, Innsbruck, Austria | Author | Major role in the acquisition of data; revised the manuscript for intellectual content |
| **Evi Holzknecht, MD** | Medical University of Innsbruck,<br>Innsbruck, Austria | Author | Major role in the acquisition of data;<br>revised the manuscript for intellectual<br>content |
| **Karel Sonka, MD, PhD** | Charles University, Prague, Czech<br>Republic | Author | Major role in the acquisition of data;<br>revised the manuscript for intellectual<br>content |
| **David Kemlink, MD,**<br>**PhD** | Charles University, Prague, Czech<br>Republic | Author | Major role in the acquisition of data;<br>revised the manuscript for intellectual<br>content |
| **Wolfgang Oertel, MD** | Philipps University, Marburg,<br>Germany | Author | Major role in the acquisition of data;<br>revised the manuscript for intellectual<br>content |
| **Annette Janzen, MD** | Philipps University, Marburg,<br>Germany |  | Major role in the acquisition of data;<br>revised the manuscript for intellectual<br>content |
| **Giuseppe Plazzi, MD** | University of Bologna, Bologna,<br>Italy | Author | Major role in the acquisition of data;<br>revised the manuscript for intellectual<br>content |
| **Elena Antelmi, MD,**<br>**PhD** | University of Bologna, Bologna,<br>Italy | Author | Major role in the acquisition of data;<br>revised the manuscript for intellectual<br>content |
| **Michela Figorilli, PhD** | University of Cagliari, Cagliari,<br>Italy | Author | Major role in the acquisition of data;<br>revised the manuscript for intellectual<br>content |
| **Monica Puligheddu,**<br>**MD** | University of Cagliari, Cagliari,<br>Italy | Author | Major role in the acquisition of data;<br>revised the manuscript for intellectual<br>content |
| **Brit Mollenhauer, MD** | University Medical Centre<br>Göttingen, Göttingen, Germany | Author | Major role in the acquisition of data;<br>revised the manuscript for intellectual<br>content |
| **Claudia Trenkwalder,**<br>**MD** | University Medical Centre<br>Göttingen, Göttingen, Germany | Author | Major role in the acquisition of data;<br>revised the manuscript for intellectual<br>content. |
| **Friederike Sixel-**<br>**Döring, MD** | University Medical Centre<br>Göttingen, Göttingen, Germany | Author | Major role in the acquisition of data;<br>revised the manuscript for intellectual<br>content |
| **Valérie Cochen De**<br>**Cock, PhD** | University Medical Centre<br>Göttingen, Göttingen, Germany | Author | Major role in the acquisition of data;<br>revised the manuscript for intellectual<br>content |
| **Christelle Charley**<br>**Monaca, MD** | University Lille North of France,<br>Lille, France | Author | Major role in the acquisition of data;<br>revised the manuscript for intellectual<br>content |
| **Anna Heidbreder, MD** | University of Münster, Münster,<br>Germany | Author | Major role in the acquisition of data;<br>revised the manuscript for intellectual<br>content |
| **Luigi Ferini-Strambi,**<br>**MD** | Università Vita-Salute San<br>Raffaele, Milan, Italy | Author | Major role in the acquisition of data;<br>revised the manuscript for intellectual<br>content |
| **Femke Dijkstra, MD** | St. Dimpna Regional Hospital,<br>Geel, Belgium | Author | Major role in the acquisition of data;<br>revised the manuscript for intellectual<br>content |
| **Mineke Viaene, MD,**<br>**PhD** | St. Dimpna Regional Hospital,<br>Geel, Belgium | Author | Major role in the acquisition of data;<br>revised the manuscript for intellectual<br>content |
| **Beatriz Abril, MD** | University Hospital of Nîmes,<br>France | Author | Major role in the acquisition of data;<br>revised the manuscript for intellectual<br>content |
| **Bradley F. Boeve, MD** | Mayo Clinic, Rochester, MN, USA | Author | Major role in the acquisition of data;<br>revised the manuscript for intellectual<br>content |
| **Ronald B. Postuma,**<br>**MD, MSc** | McGill University, QC, Canada | Author | Major role in the acquisition of data;<br>revised the manuscript for intellectual<br>content |
| **Guy A. Rouleau MD,**<br>**PhD, FRCPC** | McGill University, QC, Canada | Author | Major role in the acquisition of data;<br>revised the manuscript for intellectual<br>content |
| **Ziv Gan-Or, MD, PhD** | McGill University, QC, Canada | Author | Designed and conceptualized study; major<br>role in the acquisition of data; drafted the<br>manuscript for intellectual content |

